## Supplementary for "Patterns of testing in the extensive Danish national SARS-CoV-2 test set-up"

Index

### S1 Methods

#### SARS-CoV-2 laboratory analysis in the healthcare track

Sputum or nasopharyngeal swabs were received, each specimen marked with a barcode containing the unique national sample number, that allowed automatic linking in the laboratory system to the electronic request personal identification and requester information.

A wide range of analytical platforms and assays were used in the hospital laboratories and depending on the availability of instruments and reagents as well as the need for quick results. The platforms included rack-based in-house assays and commercial assays (e.g. Cobas 6800 from Roche, Panther from Hologic) as well as POCT instruments (e.g. Cobas LIAT from Roche, GenExpert from Cepheid). Additionally, a number of the less urgent samples, e.g. from staff screenings, were analysed at external laboratories, at universities or private companies (Novo Nordisk, Danish Technical University, Eurofins), again using their available equipment.

#### SARS-CoV-2 laboratory analysis in the community track

In general, the technological choices in the laboratory was taken in order not to interfere with the supply chain for the clinical microbiology laboratories in the healthcare track; these were in early 2020 affected by a global shortage of supplies for diagnostics. Swab samples in test tubes arrived in the lab in racks in the automation friendly 96 well configuration. Upon arrival, positive and negative controls were added to the racks. The positive controls consisted of cultivated SARS-CoV-2 virus (in house cultivated) and the negative control consisted of PBS. Racks of samples were then placed on Hamilton liquid handlers for sample registration and pre-treatment. Samples were registered by an onboard flatbed scanner, allowing 2D barcodes of the samples to be registered simultaneously. Sample pre-treatment consisted of adding 700 PBS to the swabs, agitating the samples on an on-board orbital shaker. Subsequently, 200 uL were transferred to a 96 well deep well plate. Extraction was performed on Beckman Coulter Biomek i7 liquid handlers, using the RNA advance extraction kit (Beckman Coulter, Pasadena CA). Thereafter, RT-PCR analysis was performed in using a modified version of Corman et al 2020 [1]. Briefly, 5 uL of extract was transferred to a 96 well PCR plate containing 20 uL of Luna® Universal One-Step RT-qPCR Kit (New England Biolabs, Ipswich, MA) containing primers and probes that target the E Sarbeco gene (Forward ACAGGTACGTTAATAGTTAATAGCGT, Reverse ATATTGCAGCAGTACGCACACA, Probe FAMACACTAGCCATCCTTACTGCGCTTCGBHQ1, LGC Biosearch, Lystrup Denmark). The PCR plates are sealed using a heat sealer (Thermo Scientific, Waltham MA) and were subsequently analyzed on a CFX 96 PCR thermocycler (BioRad, Hercules CA). The following cut-offs were used: CT 10 – 38, Positive; CT 38-40 inconclusive; CT >40 negative. Furthermore, all PCR curves were manually evaluated to ensure that they adhered to a sigmoid shape. Positive and negative controls must yield positive and negative results, respectively.

#### Variant Analysis of Positive Sample

Towards the end of 2020 a daily subset of positive samples was selected for whole genome sequencing (WGS). Initially, the sequencing took place at Aalborg University. Frome June 2021 a large WGS capacity was established at SSI, which eventually assumed full responsibility for sequencing of community track samples. Depending of the capabilities, the hospital laboratories either sequenced their positive samples themselves or sent them to SSI. During the Omicron wave the number of positive samples exceeded the testing capacity and the number of WGS samples was capped at 15 000 per week.

With the emergence of the Alpha (B.1.1.7) variant in early 2021, PCR based variant analysis (vPCR) was established and all positive samples were examined with vPCR. At this point, the turnaround time for initial SARS-CoV-2 testing was within 24 hours. Results from vPCR were available the day after a positive result was reported. The primary function of vPCR was to allow enhanced contact tracing of specific variants of concern. Enhanced contact tracing was performed for certain variants of concern (e.g. the Delta variant). This tracked contacts up to 3 degrees of separation. As new variants emerged, the vPCR was further developed to detect the new variants of concern. vPCRs were developed to detect Alpha (B.1.1.7), Beta (B.1.351), Delta (B.1.617.2) and Omicron (B.1.1.529) variants. The variant analysis has previously been described [2].

After analysis, all positive samples examined were stored at -80 ˚C in the Danish National Biobank (DNB) or at the local hospital. The biobanked samples were key in being able to rapidly develop and validate new vPCR methods. Samples for method validation were selected and easily retrieved from the biobank based on their WGS results. Furthermore, biobank samples were used for cultivating variant specific controls.

#### Laboratory set-up Sample logistics in the community track

The TestCenter Denmark organisation was established in April 2020. Its core was the automated, high capacity laboratory system, that was built (initially in barracks) at SSI. Swab samples were transported by car to the central laboratory, purified and PCR-analysed. As Copenhagen is located in the eastern part of the country, 10 months thereafter a twin laboratory was established in Aarhus, Jutland, in the western part of Denmark, to reduce response time and increase capacity. During the pandemic, the laboratory capacity of these two SSI-laboratories reached 200.000 daily PCR tests. The mandated performance requirement was that 80% of the samples should be reported within 24 hours and with a typical “turnaround” time, the time from taking the swab until the result was reported to the user, being between 10 to 24 hours. The laboratory facilities were expanded throughout the pandemic to keep up with demand. To maximise use of the facilities, laboratory working hours shifted from normal working hours to round the clock operation 7 days per week resulting in short turnaround times.

Mobile sampling units were also employed, though on a much smaller scale, offering added capacity during local outbreaks. Additionally, a program was set up for regular sampling at nursing homes and elder care facilities. This was performed by the municipalities that are responsible for these facilities while the testing stations were the responsibility of the authorities running the hospital system (Denmark’s healthcare system is maintained by five regional elected bodies). Thus, the community track, as organised by the TestCentre Denmark organisational structure, represented a collaboration between all three levels of administrative structures in Denmark: state (to which the SSI belongs), regional (hospitals) and municipal authorities while the regional authorities remained responsible for the healthcare track. During the start up period, the community track lab was supported by large private partners (pharma industry and management consultants and logistical support was provided by the Danish Military.

### S1 Table. Definition of heritage (as defined by Statistics Denmark)

| **Heritage** | **Definition** |
| --- | --- |
| Danish | Individuals who were born in Denmark or abroad and have at least one parent who is a Danish citizen and born in Denmark. |
| Western | Individuals with country of origin*: Nordic countries, EU countries, Andorra, Liechtenstein, Monaco, San Marino, Switzerland, United Kingdom, the Vatican City, Canada, USA, Australia or New Zealand. |
| Non-western | Individuals with country of origin*: all other countries than the countries defined above. |

*Country of origin is defined as follows:

- When neither parent is known, the country of origin is defined on the basis of the person's own information. If the person is an immigrant, it is assumed that the country of origin is equal to the country of birth. If the person is a descendant, it is assumed that the country of origin is equal to the country of citizenship.
- When only one parent is known, the country of origin is defined based on that parent’s country of birth. If this is Denmark, the country of citizenship is used.
- When both parents are known, the country of origin is defined on the basis of the mother's country of birth and country of citizenship, respectively.

### S2 Table. Unadjusted and adjusted IRR for PCR testing in the community track by sex, age, vaccination status, infection status, heritage and type of area in three study periods

|  | **February 27, 2020 to December 16, 2020 (Period I)** | | | | **December 17, 2020 to September 30, 2021 (Period II)** | | | | **November 11, 2021 to March 10, 2022 (Period III)** | | | |
| --- | --- | --- | --- | --- | --- | --- | --- | --- | --- | --- | --- | --- |
| **Sex** | **Number of tests** | **PYRS** | **Unadjusted IRR (95% CI)** | **Adjusted* IRR (95% CI)** | **Number of tests** | **PYRS** | **Unadjusted IRR (95% CI)** | **Adjusted* IRR (95% CI)** | **Number of tests** | **PYRS** | **Unadjusted IRR (95% CI)** | **Adjusted* IRR (95% CI)** |
| Female | 3,039,582 | 2,260,061 | Reference | Reference | 15,600,708 | 2,245,914 | Reference | Reference | 9,128,178 | 936,809 | Reference | Reference |
| Male | 2,502,933 | 2,286,440 | 0.83 (0.83-0.83) | 0.81 (0.81-0.82) | 12,307,701 | 2,219,523 | 0.80 (0.80-0.80) | 0.76 (0.76-0.76) | 7,193,451 | 926,134 | 0.80 (0.80-0.80) | 0.78 (0.78-0.78) |
| **Age groups (years)** |  |  |  |  |  |  |  |  |  |  |  |  |
| 2-9 | 309,956 | 392,516 | 0.46 (0.46-0.46) | 0.45 (0.45-0.45) | 1,422,968 | 382,209 | 0.54 (0.54-0.54) | 0.50 (0.50-0.50) | 1,670,302 | 158,457 | 1.01 (1.00-1.01) | 1.11 (1.11-1.12) |
| 10-19 | 942,489 | 547,511 | Reference | Reference | 3,671,104 | 534,631 | Reference | Reference | 2,308,048 | 220,403 | Reference | Reference |
| 20-29 | 1,018,508 | 624,133 | 0.95 (0.95-0.95) | 0.92 (0.91-0.92) | 4,360,788 | 610,119 | 1.04 (1-04-1.04) | 1.08 (1.08-1.08) | 2,111,702 | 254,003 | 0.79 (0.79-0.80) | 0.81 (0.81-0.81) |
| 30-39 | 762,747 | 548,601 | 0.81 (0.81-0.81) | 0.79 (0.79-0.79) | 4,286,786 | 544,116 | 1.15 (1.15-1.15) | 1.22 (1.22-1.22) | 2,355,245 | 230,021 | 0.98 (0.98-0.98) | 1.00 (0.99-1.00) |
| 40-49 | 882,686 | 605,482 | 0.85 (0.84-0.85) | 0.84 (0.84-0.84) | 5,220,315 | 588,405 | 1.29 (1.29-1.29) | 1.42 (1.42-1.42) | 2,768,654 | 239,970 | 1.10 (1.10-1.10) | 1.07 (1.07-1.08) |
| 50-59 | 836,137 | 641,084 | 0.76 (0.76-0.76) | 0.76 (0.75-0.76) | 5,125,964 | 628,200 | 1.19 (1.19-1.19) | 1.35 (1.35-1.35) | 2,596,426 | 261,865 | 0.95 (0.95-0.95) | 0.90 (0.89-0.90) |
| 60-69 | 484,912 | 530,164 | 0.53 (0.53-0.53) | 0.53 (0.53-0.53) | 2,735,916 | 521,847 | 0.76 (0.76-0.76) | 0.91 (0.90-0.91) | 1,564,498 | 218,516 | 0.68 (0.68-0.69) | 0.63 (0.63-0.63) |
| 70-79 | 248,343 | 448,605 | 0.32 (0.32-0.32) | 0.32 (0.32-0.32) | 932,586 | 445,628 | 0.30 (0.30-0.31) | 0.38 (0.38-0.39) | 705,463 | 187,858 | 0.36 (0.36-0.36) | 0.32 (0.32-0.32) |
| 80-89 | 52,347 | 176,299 | 0.17 (0.17-0.17) | 0.17 (0.17-0.17) | 140,658 | 178,567 | 0.11 (0.11-0.12) | 0.16 (0.16-0.16) | 195,844 | 77,961 | 0.24 (0.24-0.24) | 0.21 (0.21-0.21) |
| 90+ | 4,390 | 32,106 | 0.08 (0.08-0.08) | 0.08 (0.08-0.08) | 11,324 | 31,716 | 0.05 (0.05-0.05) | 0.07 (0.07-0.07) | 45,447 | 13,890 | 0.31 (0.31-0.32) | 0.27 (0.27-0.27) |
| **Vaccination status** |  |  |  |  |  |  |  |  |  |  |  |  |
| Unvaccinated or first vaccine dose | - | - | - | - | 24,656,638 | 3,298,946 | Reference | Reference | 3,188,698 | 363,424 | Reference | Reference |
| Second vaccine dose | - | - | - | - | 3,249,250 | 1,164,137 | 0.37 (0.37-0.37) | 0.40 (0.40-0.40) | 6,562,126 | 680,268 | 1.10 (1.10-1.10) | 1.18 (1.17-1.18) |
| Third vaccine dose | - | - | - | - | - | - | - | - | 6,570,805 | 819,251 | 0.91 (0.91-0.92) | 1.19 (1.19-1.19) |
| **SARS-CoV-2 infection status** |  |  |  |  |  |  |  |  |  |  |  |  |
| No infection | 5,523,841 | 4,542,922 | Reference | Reference | 27,103,688 | 4,320,890 | Reference | Reference | 15,064,692 | 1,698,544 | Reference | Reference |
| Previous infected | 18,674 | 3,579 | 4.29 (4.23-4.35) | 3.60 (3.55-3.66) | 804,721 | 144,547 | 0.89 (0.89-0.89) | 0.88 (0.88-0.89) | 1,256,937 | 164,400 | 0.86 (0.86-0.86) | 0.83 (0.83-0.83) |
| **Heritage** |  |  |  |  |  |  |  |  |  |  |  |  |
| Danish | 4,631,767 | 3,831,340 | Reference | Reference | 24,403,264 | 3,763,667 | Reference | Reference | 1,4262,429 | 1,564,242 | Reference | Reference |
| Non-west | 541,180 | 410,428 | 1.09 (1.09-1.09) | 0.90 (0.90-0.90) | 1,936,982 | 407,781 | 0.73 (0.73-0.73) | 0.60 (0.60-0.60) | 1,172,122 | 172,830 | 0.74 (0.74-0.75) | 0.69 (0.69-0.69) |
| West | 365,905 | 287,170 | 1.05 (1.05-1.06) | 0.90 (0.90-0.91) | 1,551,769 | 289,432 | 0.83 (0.83-0.83) | 0.69 (0.69-0.69) | 883,992 | 125,222 | 0.77 (0.77-0.78) | 0.73 (0.74-0.74) |
| Unknown | 3,663 | 17,562 | 0.17 (0.17-0.18) | 0.21 (0.20-0.21) | 16,394 | 4,558 | 0.55 (0.55-0.56) | 0.53 (0.53-0.54) | 3,086 | 649 | 0.52 (0.50-0.54) | 0.52 (0.50-0.54) |
| **Type of area (municipality)** |  |  |  |  |  |  |  |  |  |  |  |  |
| Capital Municipalities | 1,921,590 | 1,274,950 | Reference | Reference | 8,101,930 | 1,256,438 | Reference | Reference | 4,566,901 | 526,660 | Reference | Reference |
| Commuter Municipalities | 666,899 | 706,691 | 0.62 (0.62-0.63) | 0.66 (0.65-0.66) | 4,035,668 | 692,702 | 0.90 (0.90-0.90) | 0.90 (0.90-0.91) | 2,424,152 | 288,009 | 0.97 (0.97-0.97) | 0.97 (0.97-0.97) |
| Metropolitan Municipalities | 804,578 | 619,500 | 0.86 (0.86-0.86) | 0.85 (0.85-0.85) | 4,099,980 | 609,055 | 1.04 (1.04-1.05) | 1.02 (1.02-1.02) | 2,125,768 | 254,777 | 0.96 (0.96-0.96) | 0.94 (0.94-0.94) |
| Provincial Municipalities | 1,099,508 | 1,028,583 | 0.71 (0.71-0.71) | 0.73 (0.73-0.73) | 6,327,699 | 1,009,594 | 0.97 (0.97-0.97) | 0.96 (0.96-0.97) | 376,7397 | 420,573 | 1.03 (1.03-1.03) | 1.02 (1.02-1.02) |
| Rural Municipalities | 1,049,940 | 916,778 | 0.76 (0.76-0.76) | 0.81 (0.81-0.81) | 5,343,132 | 897,650 | 0.92 (0.92-0.92) | 0.94 (0.94-0.94) | 3,437,411 | 372,925 | 1.06 (1.06-1.06) | 1.09 (1.08-1.09) |

*Adjusted for age group, sex, vaccination status, SARS-CoV-2 infection status, heritage, and type of area
